# Prediction of Gastrointestinal Bleeding Hospitalization in Hemodialysis

**DOI:** 10.1101/2023.01.31.23285098

**Authors:** John W. Larkin, Suman Lama, Sheetal Chaudhuri, Joanna Willetts, Anke C. Winter, Yue Jiao, Manuela Stauss-Grabo, Len A. Usvyat, Jeffrey L. Hymes, Franklin W. Maddux, David C. Wheeler, Peter Stenvinkel, Jürgen Floege, the INSPIRE Core Group

**Author notes:** **Corresponding Author:** John Larkin, PhD, Fresenius Medical Care, Global Medical Office, 920 Winter Street, Waltham, MA 02451, United States.

## Abstract

Gastrointestinal bleeding (GIB) is a clinical challenge in kidney failure. The INSPIRE group assessed if machine learning could assist with determining a hemodialysis (HD) patient’s 180-day GIB hospitalization risk. Model was developed using adult HD patient data from United States (2017-2020). Patient data was randomly split (50% training, 30% validation, and 20% testing). HD treatments ≤ 180 days before GIB hospitalization were classified as positive observations, and others were negative observations. Datasets were randomly sampled to build an XGBoost model that considered 386 exposures initially and was refined to the top 50 exposures. Unseen testing dataset was used to determine final model performance. Incidence of 180-day GIB hospitalization was 1.18% in the HD population (n=451,579), and 1.16% among patients in the testing dataset (n=27,991). Model showed an area under the curve=0.69, sensitivity=57.9%, specificity=68.9%, accuracy=68.8% and balanced accuracy=63.4%. Exposures with largest effect size per Shapley values were older age (group mean GIB event=68.2 years vs no GIB event=63.4 years), shorter days since last all-cause hospital admission (group mean GIB event=203.2 days vs no GIB event=253.2 days), and higher serum 25-hydroxy (OH) vitamin D levels from most recent lab (group mean GIB event=33.4 ng/mL vs no GIB event=30.5 ng/mL). Other important predictors included lower hemoglobin and iron indices, longer dialysis vintage, and proton pump inhibitor use. Model appears suitable for early detection of GIB event risk in HD, yet prospective testing is needed. The association between higher 25OH vitamin D and GIB events was unexpected and warrants investigation.

## Introduction

**IN**itiative**S** on advancing **P**atients’ outcomes In **RE**nal disease (**INSPIRE**) is an academia and industry collaboration set forth to identify critical investigations/models needed to advance the practice of medicine in nephrology. At the inaugural INSPIRE meeting, a Core Group of nephrology professionals chose major gastrointestinal bleeding (GIB) events as a top priority. The consensus was severe bleeding events represent potentially preventable complications that occur more frequently in people with kidney disease as compared to the general population.^1-6^

Major bleeding events have a 2% to 6% incidence per year among dialysis patients.^7-9^ Bleeding events differ by modality, with higher rates seen in hemodialysis (HD) compared to peritoneal dialysis (PD).^10^ Most bleeding events in dialysis patients are due to a gastrointestinal bleed (GIB), with about 20% requiring hospitalization.^7, 11^ The incidence of GIB hospitalizations has been increasing over time in the dialysis population.^11^ Dialysis patients who experienced a prior GIB have an 90% higher risk of death, and this risk increases with each subsequent GIB event.^11^

Although bleeding risk scores have been developed for various patient populations (e.g., GBS,^12^ HAS-BLED,^13^ ATRIA,^14^ HEMORR2HAGES,^15^ ORBIT^16^), they have poor performance in dialysis patients. ^8, 17, 18^ Machine learning methods have been evaluated as a way to help identify dialysis patients at higher risk for an all-cause bleeding event, yet have so far had inadequate performance to improve detection.^8^ The inability to identify a dialysis patient’s risk for an ensuing bleeding event might be due to the classification for all-cause events, rather than specific types of bleeding events that can have distinct clinical characteristics defining the condition. The INSPIRE Core Group aimed to develop a machine learning model to determine if artificial intelligence-based methods may be able to provide suitable identification of a HD patient’s risk for hospitalization due to a GIB event.

## Methods

### Patient Population

To identify a unique patient’s risk of a GIB hospitalization in the next 180 days, we utilized real-world retrospective data from adults (age ≥18 years) who received ≥1 outpatient HD treatment at an integrated kidney disease company with approximately 2,500 dialysis centers (Fresenius Kidney Care, Waltham, United States) during 01-Jan-2017 through 31-Dec-2020.

This project was reviewed and approved by New England Independent Review Board (Needham Heights, MA, United States; Work Oder# 1-1502098-1). It was determined by the Independent Review Board that this analysis was exempt due to deidentification of data and consent was not required per title 45 Code of Federal Regulations part 46.104(d)(4) in the United States. The analysis adhered to the Declaration of Helsinki.

### Outcome and Predictor Variables

The outcome (dependent variable) was defined as a GIB event requiring hospitalization as determined from the discharge diagnosis ICD10 codes: K22.6, K25.0, K25.2, K25.4, K25.6, K26.0, K26.2, K26.4, K26.6, K27.0, K27.2, K27.4, K27.6, K28.0, K28.2, K28.4, K28.6, K29.0, K62.5, K66.1, K92.0, K92.1, K92.2. The at-risk exposure time for the prediction of the outcome was investigated and chosen to be within 180 days before the prediction date. The prediction date was based on each HD treatment observation date across the study period.

We used a data driven approach to investigate differing exposures (independent variables) considering both clinical factors based on *a priori* assumptions, as well as an array of various parameters routinely captured in dialysis care. We chose this approach since the risk factors for an ensuing GIB hospitalization have not been clearly defined in the dialysis population. Also, the machine learning model we utilized had a favorable attribute of being able to select the exposures with the most importance to an outcome prediction in an individualized manner for each patient.^19^ The exposure variables included an array of demographics, comorbidities, environmental attributes, laboratories, HD treatment data, medications, and prior events (**Supplementary Table 1**). For each unique parameter (n=145) we included the most recent value/status as of the prediction date, and for continuous data, also included the mean values in the prior 7, 30, 90, and/or 180 days as it was appropriate considering data availability (**Figure 1**).

**Figure 1:**
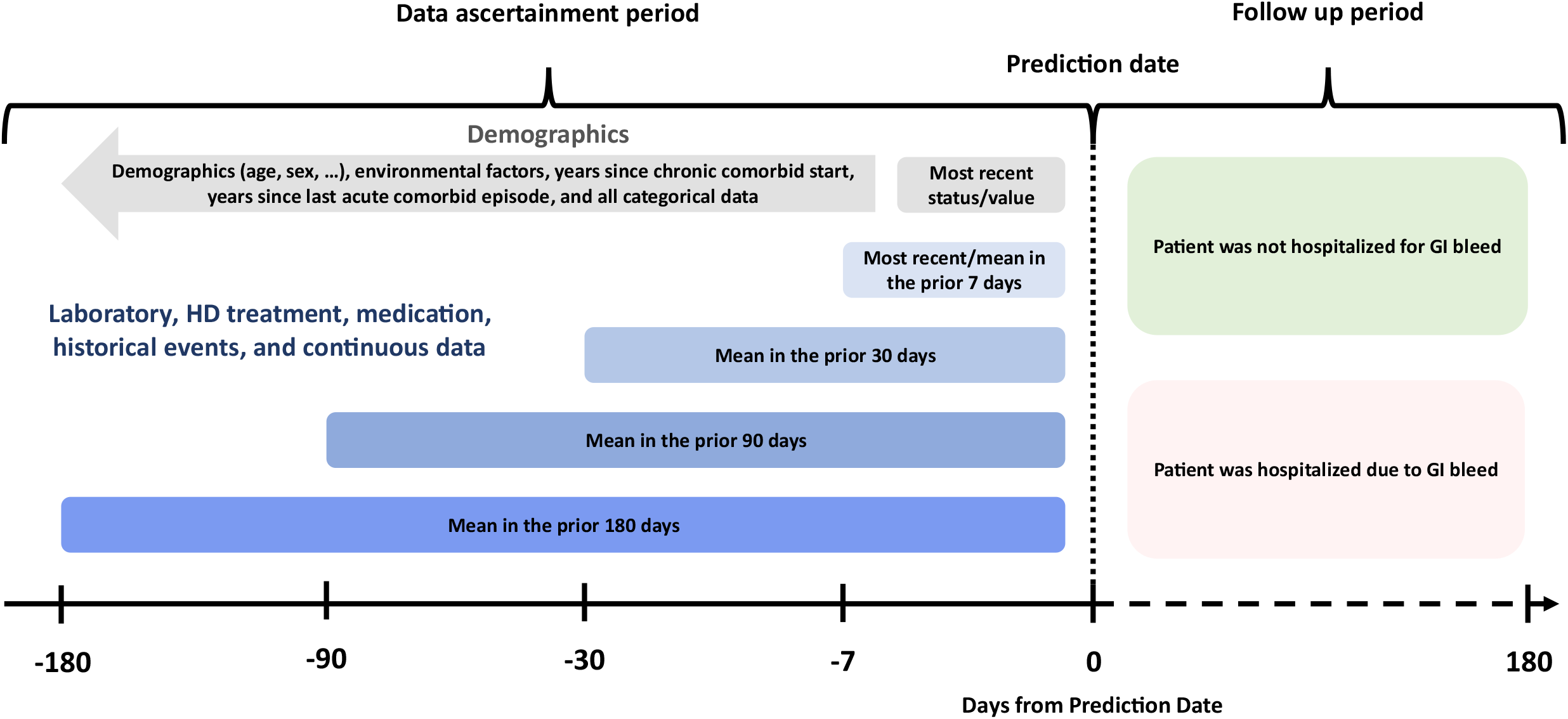
Data ascertainment and outcome follow up timeframes.

Erythropoietin stimulating agent (ESA) doses were converted into continuous erythropoiesis receptor activator (CERA) equivalent units for analysis using previously established ratios.^20^ Intravenous vitamin D medications were converted to doxercalciferol equivalent units for analysis using a conversion ratio of 1:1.54 from paricalcitol (i.e. 65% of the paricalcitol dose) and a conversion ratio of 1:1.375 from calcitriol (i.e. 73% of the calcitriol dose).^21, 22^ Proton pump inhibitor (PPI), anticoagulant, and antiplatelet medication classes were defined using the United States Food and Drug Administration’s National Drug Code Directory.^23^

Overall, the model assessed 386 exposure variables for individualized predictions (both unique and calculated variables). For the final model developed and evaluated in this report, we refined the exposure variables using a data driven approach that considered the top 50 exposure variables determined to exhibit the highest importance in the initial model. This method was selected to optimize exploratory assessments, while using a practical number of exposure variables that typically have the most meaning for making the prediction.

### Model Development

We organized data for model development by splitting patient records and sampling observations. Data from distinct patients was randomly split into a training (50% of patients), validation (30% of patients), or testing (20% of patients) dataset. In these datasets, each HD treatment observation within 180 days before a GIB hospitalization was classified as a positive observation (i.e., experienced a GIB event in next 180 days). All other HD treatment observations were classified as a negative observation (i.e., did not have GIB event in next 180 days). We then randomly selected a subset of observations to be used for model development, considering samples from the positive and negative observations within these three datasets (**Supplementary Figure 1**). This sampling considered an equivalent number of observations with positive and negative GIB events in the training dataset, and an incidence that matched the overall population in the validation and testing datasets.

We used Python version 3.7.7 (Python Software Foundation, Delaware, United States) with the XGBoost package for constructing the machine learning model.^24^ For model development, XGBoost first used the training dataset and constructed decision trees for the exposure variables in every possible combination and established a series of thresholds, splitting variables to maximize the information gain. This ensemble of decision trees was constructed iteratively, and new decision trees were added to predict prior errors. The model was inherently able to manage missing values without imputation by including the variables presence for each patient when establishing variable splits. The model was next run on the validation dataset, where it was adjusted and tuned until no further improvements in performance were achieved. This model was used to identify the importance of the predictors and establish the top 50 exposure variables to be considered in the final model. The model construction then started over again including only the top 50 exposure variables, and the training and validation steps were repeated in the same manner. This ensemble of decision trees produced the final model that was used to assess the model’s performance on the unseen data in a testing dataset.

### Importance of Predictor Variables

The importance of exposure variables was determined using Shapley (SHAP) values ^25, 26^ computed using the SHAP python package ^27, 28^. SHAP values determined each exposure variable’s importance by assessing the effect size of each unique variable, and the overall combination of variables, on the prediction. The levels of importance (i.e., the effect size) found by SHAP values were used for the selection of the top 50 predictor variables to be included in the final model, and show the relative meaningfulness of each top predictor.

The logic behind the calculation of SHAP values included a measurement of impact (positive or negative value) for each variable at each observation for each individual patient’s prediction. SHAP methods withheld and included individual variables in all combinations to compute the mean values for attributing the importance for each exposure variable. SHAP values show the effect size as log odds (i.e., the logarithm of the odds ratio) and represent non-linear additive explanations for variable importance. SHAP values for each variable were summed for each patient and can be converted from log odds to probability for each patient’s individualized prediction. Basically, the SHAP value shows the effect size and the direction of the effect (positive log odds value show higher risk and negative value show lower risk) for each exposure variable from each unique patient’s individualized prediction. The top predictors for each patient include those with the greatest effect size/importance for that patient’s prediction.

The overall effect size/importance of the exposure variables on the patient population was determined using an absolute value (non-negative value) of the SHAP value for each exposure variable, taking the mean value for all distinct patients. Therefore, the overall population mean SHAP value shows the effect size for each exposure variable based on the predictions from all patients. The mean SHAP value was used for ranking the overall risk attributable to each exposure variable and defined the top predictors of the outcome of a GIB hospitalization in the next 180 days.

### Assessment of Model Performance

Model performance was measured by the area under the receiver operating characteristic curve (AUROC), sensitivity, specificity, accuracy, and balanced accuracy; these were assessed in the training, validation, and testing datasets used for model development. The final performance assessment was considered for only the unseen testing dataset considering a cutoff threshold of 0.50. The details of the performance metrics are denoted below.

AUROC: this metric shows the rate of true and false positives classified by the prediction model across probability thresholds. True and false positives and negatives are defined in **Supplementary Table 2**.

Sensitivity (also known as recall): this metric shows the rate of true positives classified by the model at a specified threshold and was calculated as follows:

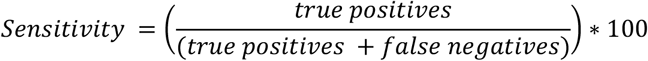

Specificity: this metric shows the rate of true negatives classified by the model at a specified threshold and was calculated as follows:

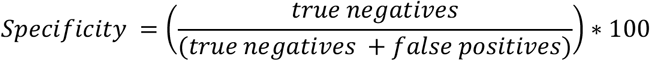

Accuracy: This metric shows the rate of true positives and true negatives classified by the model at a specified threshold (i.e., the fraction of correct predictions) and was calculated as follows:

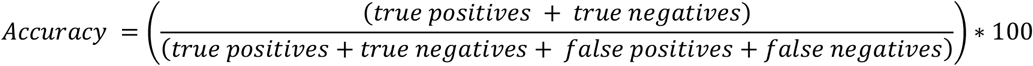

Balanced accuracy: This metric shows the mean of the sensitivity and specificity of the model and was calculated as follows:

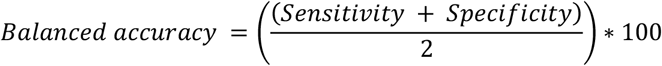

The metrics for AUROC, sensitivity, specificity, accuracy, and balanced accuracy computed scores on a scale of 0 (lowest) to 1 (highest). Sensitivity, specificity, accuracy, and balanced accuracy are presented as a percentage. As an example, a model performing at random chance would have an AUROC = 0.5 and a balanced accuracy = 50%.

## Results

### Patient Population Characteristics

We considered a patient population of 451,579 adults treated by HD during 2017 throughout 2020. The characteristics of the patient population are shown in **Table 1** for those who experienced ≥1 GIB hospitalization or no GIB hospitalization. The incidence of GIB hospitalization within 180 days of a given HD treatment was 1.18% (1,249,108/105,838,571 observations) in the patient population.

**TABLE 1:** CHARACTERISTICS OF THE HD PATIENT POPULATION, RANDOM SUBSET OF PATIENTS, AND TEST DATASET OF PATIENTS.

| Parameter | GIB Admission<br>Mean±SD or % |  |  | No GIB Admission<br>Mean±SD or % |  |  |
| --- | --- | --- | --- | --- | --- | --- |
|  | Population | Subset | Test Dataset | Patient Population | Subset | Test Dataset |
| Patient n | 28644 | 4107 | 436 | 422935 | 207184 | 27555 |
| Observation n | 1,249,108 | 98,244 | 468 | 104,589,463 | 178,725 | 39,720 |
| Age (years) | 67.3 ± 13.0 | 68.2 ± 12.7 | 68.2 ± 12.6 | 62.8 ± 14.6 | 63.3 ± 14.2 | 63.4 ± 14.2 |
| Male | 55% | 55% | 51% | 58% | 57% | 57% |
| White Race | 48% | 47% | 47% | 44% | 47% | 47% |
| Black Race | 28% | 31% | 31% | 22% | 30% | 30% |
| Asian Race | 20% | 17% | 18% | 30% | 18% | 19% |
| Other Race | 3% | 3% | 3% | 2% | 3% | 3% |
| Unknown Race | 1% | 1% | 1% | 1% | 2% | 2% |
| Hispanic Ethnicity | 67% | 68% | 67% | 56% | 65% | 65% |
| Not Hispanic Ethnicity | 9% | 10% | 11% | 10% | 13% | 13% |
| Unknown Ethnicity | 24% | 22% | 22% | 34% | 23% | 23% |
| Dialysis vintage (years) | 3.9 ± 4.1 | 4.5 ± 4.2 | 4.4 ± 4.0 | 2.3 ± 3.6 | 4.2 ± 4.1 | 4.2 ± 4.1 |
| Catheter HD access | 28% | 19% | 20% | 43% | 18% | 18% |
| Arteriovenous HD access | 70% | 81% | 80% | 46% | 81% | 82% |
| Diabetes | 39% | 40% | 41% | 30% | 39% | 40% |
| Ischemic heart disease | 24% | 25% | 19% | 15% | 19% | 19% |
| GIB (comorbidity) | 2% | 3% | 3% | 0.4% | 1% | 0.8% |
| Self-reported energy level<sup>†</sup> | 45.3 ± 27.8 | 45.5 ± 27.5 | 46.8 ± 26.8 | 46.3 ± 27.3 | 50.2 ± 27.8 | 50.1 ± 27.8 |

<sup>†</sup> KDQOL-36 score for question 10: “Did you have a lot of energy?”
GIB: gastrointestinal bleed; HD: hemodialysis

We split the patient population into three groups, randomly assigning each distinct patient’s data into a training (n=225,793), validation (n=135,490), or testing (n=90,296) dataset. We then randomly selected subset of observations for the training (patient n=76,441), validation (patient n=31,288), and testing (patient n=27,991) datasets used to construct the model. This sampling considered an equivalent number of observations with positive and negative GIB events in the training dataset, and an incidence that matched the overall population in the validation and testing datasets. The patient characteristics in the subset of data used were reasonably consistent with the overall patient population, albeit there were some small differences in parameters after random splitting/sampling (**Table 1**).

### Model Performance

The machine learning prediction model was developed using distinct groups of patients who had their data randomly assigned to training and validation datasets. Initially, the model used 145 distinct exposure variables and considered the most recent values to the prediction date, as well as mean values for continuous variables in prior 7, 30, 90, and 180 days (i.e., 386 distinct & calculated exposure variables; **Supplementary Table 1**). The final model was refined to include only the top 50 variables that showed the highest importance for outcome prediction (**Table 2**). The performance of the prediction model was evaluated on the testing dataset, which was unseen by the model during development. The prediction model was found to have suitable performance in the classification of patients at a higher risk of being hospitalized for a GIB in the following 180 days (**Table 3**). In the testing dataset, the model showed an AUROC of 0.69, a sensitivity of 57.9%, a specificity of 68.9%, and an accuracy of 68.8%.

**TABLE 2:** PREDICTORS OF 180-DAY GI BLEED HOSPITALIZATION IN THE TEST DATASET.

| Parameter | Mean SHAP value | GIB Admission Mean (SD) or % | No GIB Admission Mean (SD) or % |
| --- | --- | --- | --- |
| Patient n (test dataset) | N/A | 436 | 27,555 |
| Observation n (test dataset) | N/A | 468 | 39,720 |
| Age (years) | 0.071 | 68.2 (12.6) | 63.4 (14.2) |
| Days since start of last all-cause hospitalization | 0.045 | 203.2 (245.7) | 253.2 (268.7) |
| 25OH Vitamin D (ng/mL): last lab | 0.044 | 33.4 (17.0) | 30.5 (16.0) |
| 25OH Vitamin D (ng/mL): mean 180 days | 0.025 | 32.8 (15.6) | 30.4 (15.9) |
| Hgb (g/dL): last lab | 0.024 | 10.3 (1.4) | 10.8 (1.3) |
| Ferritin (ng/mL): mean 180 days | 0.021 | 948.7 (529.8) | 1005.3 (553.3) |
| TSAT (%): mean 180 days | 0.020 | 31.0 (9.8) | 32.8 (9.9) |
| Hgb (g/dL): mean 180 days | 0.020 | 10.3 (1.0) | 10.7 (1.0) |
| Dialysis vintage (years) | 0.017 | 4.4 (4.0) | 4.2 (4.1) |
| EDW: mean 7 days | 0.013 | 78.3 (22.4) | 82.8 (23.5) |
| Albumin (g/dL): last lab | 0.012 | 3.7 (0.5) | 3.8 (0.4) |
| Hgb (g/dL): mean 90 days | 0.012 | 10.4 (1.1) | 10.7 (1.1) |
| Albumin (g/dL): mean 180 days | 0.012 | 3.7 (0.4) | 3.8 (0.4) |
| Vitamin B12 (pg/mL): last lab | 0.011 | 786.2 (411.4) | 716.4 (367.6) |
| Vitamin B12 (pg/mL): mean 180 days | 0.011 | 773.6 (401.9) | 719.9 (363.9) |
| QB (mL/min): mean 90 days | 0.011 | 406.4 (47.8) | 411.6 (49.7) |
| Lymphocytes (%): mean 180 days | 0.010 | 18.6 (7.1) | 20.1 (7.3) |
| Ferritin (ng/mL): last lab | 0.010 | 953.7 (538.3) | 1026.6 (566.9) |
| Chloride (mEq/L): mean 90 days | 0.010 | 98.7 (3.7) | 99.0 (3.6) |
| Hgb (g/dL): mean 7 days | 0.010 | 10.3 (1.4) | 10.8 (1.3) |
| Pre-HD DBP: mean 90 days | 0.010 | 74.1 (12.5) | 77.1 (11.9) |
| Bicarb (mEq/L): mean 180 days | 0.009 | 24.2 (2.4) | 24.0 (2.3) |
| Platelets (1000/mcL): mean 180 days | 0.009 | 192.9 (69.8) | 199.8 (67.5) |
| Lymphocytes (%): mean 90 days | 0.009 | 18.4 (7.4) | 20.1 (7.5) |
| TSAT (%): mean 90 days | 0.009 | 31.0 (11.1) | 33.0 (11.1) |
| PPI Use | 0.009 | 23.3% | 20.5% |
| EDW: last HD | 0.008 | 78.2 (22.3) | 82.8 (23.5) |
| Hgb A1C: mean 180 days | 0.008 | 6.2 (1.3) | 6.6 (1.5) |
| BUN to Cr ratio: mean 90 days | 0.008 | 7.6 (3.1) | 7.4 (3.1) |
| WBC (1000/mcL): mean 180 days | 0.007 | 6.8 (2.3) | 6.9 (2.6) |
| Monocytes (%): mean 180 days | 0.007 | 6.5 (1.6) | 6.3 (1.6) |
| Prothrombin Time (seconds): last lab | 0.007 | 24.5 (9.1) | 21.2 (8.1) |
| CERA dose (mcg): mean 90 days | 0.007 | 156.8 (147.2) | 89.8 (71.8) |
| Non-Hispanic Ethnicity | 0.006 | 67.3% | 64.6% |
| Platelets (1000/mcL): mean 90 days | 0.006 | 191.5 (73.9) | 199 (69.3) |
| Post-HD stand SBP: mean 30 days | 0.006 | 137.2 (19.9) | 135.3 (18.6) |
| Hgb A1C: last lab | 0.006 | 6.2 (1.4) | 6.6 (1.6) |
| Post-HD pulse: mean 7 days | 0.006 | 75.8 (11.2) | 76.5 (11.1) |
| Pre-HD stand DBP: mean 90 days | 0.005 | 76.4 (13.2) | 78.3 (12.6) |
| HD session stopped against med advice: mean % 90 days | 0.005 | 2.5% | 1.8% |
| IV iron sucrose: mean 90 days | 0.005 | 85.0 (24.2) | 75.8 (26.6) |
| Intact PTH: mean 180 days | 0.005 | 439.6 (346.2) | 467.6 (336.4) |
| Basophils (%): mean 90 days | 0.005 | 0.7 (0.4) | 0.7 (0.4) |
| Lymphocytes (%): mean 30 days | 0.005 | 18.5 (8.2) | 20.1 (8.1) |
| KECN: mean 90 days | 0.005 | 253.7 (26.6) | 254.3 (27.6) |
| Ferritin (ng/mL): mean 90 days | 0.005 | 943.9 (525.8) | 1018.3 (551.8) |
| HD session stopped unexpectedly: mean % 90 days | 0.004 | 8.5% | 7.3% |
| BUN to Cr ratio: last lab | 0.004 | 7.7 (3.4) | 7.4 (3.2) |
| Creatinine (mg/dL): last lab | 0.003 | 7.9 (2.7) | 8.4 (3.0) |
| Length of stay (days) for last hospitalization | 0.003 | 5.0 (6.0) | 4.6 (9.2) |

GIB: gastrointestinal bleed; EDW: estimated dry weight; CERA: continuous erythropoiesis receptor activator; Bicarb: bicarbonate; IV: intravenous; Hgb: hemoglobin; TSAT: transferrin saturation; PPI: proton pump inhibitor; Hgb A1C: hemoglobin A1C; QB: blood flow rate; BUN to Cr ratio: blood urea nitrogen to creatinine ratio; PTH: parathyroid hormone; WBC: white blood cells; HD: hemodialysis; SD: standard deviation; Stand: standing; SBP: systolic blood pressure; DBP: diastolic blood pressure; KECN: effective clearance of sodium

**TABLE 3:** MODEL PERFORMANCE IN PREDICTING 180-DAY GI BLEED HOSPITALIZATION RISK.

| **Model Dataset** | **Training** | **Validation** | **Testing** |
| --- | --- | --- | --- |
| **Patient n** | 76,441 | 31,288 | 27,991 |
| **Observation n** | 196,589 | 40,192 | 40,188 |
| **Incidence of GI bleed hospitalization observations per 180 days** | 49.5% | 1.17% | 1.16% |
| **AUROC** | 0.738 | 0.714 | 0.691 |
| **Sensitivity (Recall)** | 65.4% | 58.3% | 57.9% |
| **Specificity** | 69.1% | 69.9% | 68.9% |
| **Accuracy** | 67.3% | 69.7% | 68.8% |
| **Balanced accuracy** | 67.2% | 64.1% | 63.4% |

GI: gastrointestinal; AUROC: area under the receiver operating characteristic curve

### Predictors of GIB Hospitalization

SHAP values were computed to determine the effect size for each variable in the prediction. In the unseen data in the testing dataset, the top three predictors of a GIB hospitalization in the next 180 days were older age (group mean GIB event = 68.2 years vs no GIB event = 63.4 years), shorter days since the last all-cause hospital admission (group mean GIB event = 203.2 days vs no GIB event = 253.2 days), and surprisingly, higher total serum 25-hydroxy (OH) vitamin D levels from the most recent lab (group mean GIB event = 33.4 ng/mL vs no GIB event = 30.5 ng/mL). Other top predictors included lower levels of hemoglobin and iron indices, longer dialysis vintage, lower estimated dry weight, and lower albumin levels. Albeit the 26^th^ most important predictor, we found use of a PPI was a risk factor for GIB hospitalization. Other medications in the top 50 predictors were higher ESA dose and higher IV iron dose. No other in-center or home medications were among the top 50 predictors (e.g., heparin, anticoagulants, antiplatelets, active vitamin D analogs) and were not included in the final model since they did not exhibit a great enough effect size. Nonetheless, a longer prothrombin time was the 32^nd^ most important predictor and represents the anticipated connection between anticoagulation and GIB events.

**Figure 2** shows the top 25 predictors with the greatest effect on classification of 180-day risk for a GIB hospitalization in the testing dataset. The bar chart (left panel) shows the mean absolute SHAP value (a non-negative/absolute value), which is the non-linear magnitude of the effect size for each variable in log odds. The top predictors are shown in descending order. The SHAP value plot (right panel) shows additional information on the direction as well as the magnitude of the effect size for each exposure variable from each patient’s individualized prediction. The SHAP value assigned to each dot represents the value from each individual patient for that specific exposure variable. The position of the dot on the x-axis corresponds to the effect size for that specific patient, either positive (showing more risk) or negative (showing more protection). The color each dot corresponds to the value of the exposure variable is for that specific patient. Dots with warmer colors show a higher value for a specific patient, while dots with cooler colors show a smaller value for a specific patient, and gray dots show a missing value for a specific patient.

To provide an example, the mean SHAP value for the top predictor of age found this exposure has the greatest effect size in the prediction of a GIB hospitalization in the next 180 days overall considering the entire group evaluated. The SHAP value plot further found the dots became warmer colors with more positive SHAP values (showing more risk with older ages) and the dots became cooler colors with more negative SHAP values (showing more protective effects with younger ages). Albeit age was found to contribute the most to the risk of a GIB hospitalization, there were many exposures that had a large effect size for specific patients, and this can be seen by the log odds values and the distributions of risks in SHAP value plots that show the results from individual predictions.

**Figure 2:**
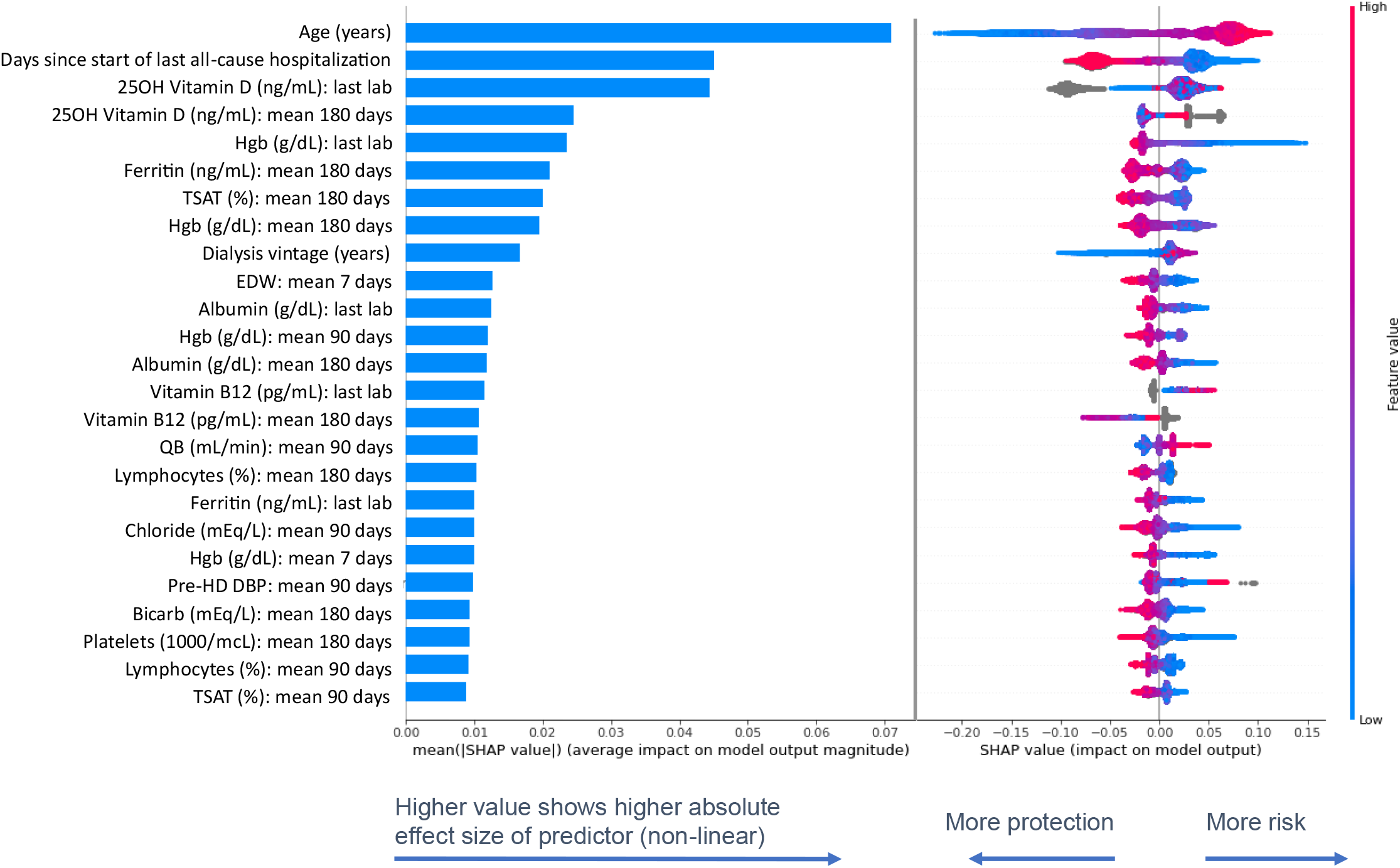
Top 25 predictors of 180-day GIB hospitalization in descending order. Bar plot on the left panel show the mean absolute SHAP values that estimate the average effect size of each exposure variable’s contribution to predicting the outcome on the x-axis (calculated from the average absolute value for all patients). SHAP value plots in the right panels show the size and direction (more positive=higher risk or more negative=lower risk/more protection of each variable’s influence on the outcome for each unique patient on the x-axis, with warmer colors representing higher observed values for that measurement, cooler colors indicating lower values for that measurement, and gray representing a missing value for that measurement. SHAP values are presented in the unit of log odds (i.e. logarithm of the odds ratio). GIB: gastrointestinal bleed; EDW: estimated dry weight; Bicarb: bicarbonate; Hgb; hemoglobin; TSAT: transferrin saturation; QB: blood flow rate; HD: hemodialysis; DBP: diastolic blood pressure.

## Discussion

Major GIB events are potentially avoidable, yet underrecognized in the HD population. To improve the ability for early detection, we developed a machine learning model to identify a HD patient’s 180-day risk for a GIB hospitalization. Model showed suitable performance and found the most important risk factors for GIB hospitalization in the patient population were older age, shorter days since the last all-cause hospitalization, and higher total 25OH vitamin D levels from the most recent lab. Albeit many of the top predictors have been previously suggested to be associated with GIB risk in HD,^11, 29^ the strong association between higher total 25OH vitamin D levels and GIB events was unexpected and warrants further investigation.

We found a 1.2% incidence of 180-day GIB hospitalization. This is consistent with other reports in the literature that show a 2% to 6% incidence per year.^7-9^ Despite a low incidence, experiencing a GIB hospitalization can increase the risk of death by 90% in a kidney failure patient,^11^ emphasizing the need to enhance early detection and treatment. GIB can be detected by fecal occult blood tests and endoscopy.^30-32^ However, there is little information to guide the use and frequency of fecal occult blood screening in the dialysis population and early detection of a suspected GIB is largely dependent on a timely referral to a gastroenterologist. Many GIBs can be effectively managed by pharmaceutical regimens or treated during screening procedures, with about 40% of upper GIBs being treated in an outpatient setting.^33^ Our model found PPI use was a meaningful predictor of a major GIB event, which is likely an observation by indication. Nonetheless, it shows some preventative measures taken among those with known gastroesophageal complications. A recent study of >200,000 hospitalized patients showed kidney failure patients had lower endoscopy rates and higher mortality rates than matched patients without kidney failure.^4^ Furthermore, this study showed kidney failure patients with a major GIB who had an endoscopy exhibited lower mortality rates than those who did not receive an endoscopy. This supports the potential benefits of endoscopy for diagnostic evaluation and treatment as appropriate. Notably, kidney failure itself is a significant risk factor for GIB.^5, 29, 34^ A study of dialysis patients who received an endoscopy during kidney transplant evaluation showed >60% of patients had abnormal endoscopic findings.^35^

There are traditional and machine learning risk models available for guiding treatment and prognosis of GIB among patients presenting to the emergency department/hospital. Despite this, these models often include kidney failure and/or markers altered in kidney disease as inputs, and thus can yield convoluted insights in the dialysis population.^36-40^ The appropriate identification and risk classification in patients with kidney failure remains a clinical challenge. The Glasgow Blatchford score (GBS) has been evaluated for predicting the need for admission and endoscopic intervention in kidney failure patients presenting to the hospital with a suspected GIB; this model was found to have reasonable performance (AUROC=0.63, sensitivity=81.2%, and specificity=42.3%) with a GBS cutoff score of ≥14.^34^ However, in comparison, a GBS cutoff score of >0 (zero) is considered appropriate to define the need for admission and endoscopy in patients without kidney failure.^12^ To our knowledge, there are presently no GIB risk prediction models specific to the outpatient kidney failure population. Nonetheless, all-cause bleeding risk models have been tested in kidney failure patients, yet none have had suitable performance to be considered in care.^8, 17, 18^ Rather than using the outcome of all-cause bleeding, we focused on the most frequent class of bleeding events and were able to build a model with reasonable performance.

We identified an unexpected and potentially important association between higher total serum 25OH vitamin D levels and major GIB. This observation is consistent with findings in warfarin users without known kidney disease who have a higher GIB risk when total serum 25OH vitamin D levels are in the range of 30-100 ng/mL versus all other levels.^41^ KDOQI guidelines suggest total serum 25OH vitamin D levels should be maintained at ≥30 ng/mL in kidney disease.^22^ Despite this, the United States Food and Nutrition Board recommends avoiding total serum 25OH vitamin D levels >50 ng/mL and suggests careful considerations for levels 30 to 49 ng/mL since they are associated with higher rates of mortality, cardiovascular events, falls/fractures and cancer.^42^ KDIGO guidelines state further research is needed to determine the benefits or risks associated with vitamin D analogs.^43^ A preliminary investigation of this signal by the INSPIRE Core Group presented as an abstract found unadjusted GIB event rates were qualitatively higher among HD patients with total serum 25OH vitamin D levels >30 ng/mL, and the highest at levels of ≥50 to <60 ng/mL.^44^ Further analyses are needed to confirm this observation.

There is a growing body of evidence suggesting both anticoagulant and antithrombotic actions of serum vitamin D levels and use of vitamin D derivatives.^45^ For instance, in vitro 1,25-dihydroxyvitamin D induces the secretion of tissue plasminogen activator from rat heart cells,^46^ down-regulates the expression of plasminogen activator inhibitor 1 in rat osteoblast cells ^47^ and human breast cancer cells,^48^ and down-regulates the expression of tissue factors in human leukemia cells.^49^ Vitamin D may also play a role in impairment of platelet aggregation with vitamin D receptor knockout mice exhibiting enhanced platelet aggregation.^50^ Consistent with our observation of a potential anticoagulant effect of vitamin D, high total 25OH vitamin D levels in the general population have been associated with a reduced venous thromboembolism risk,^51^ and use of 1,25-dihydroxyvitamin D is associated with a decreased incidence of deep vein thrombosis among prostate cancer patients.^52^

Although we were able to construct a model that may be suited for further evaluation, there are several limitations to be considered. We used a subset of historic data for model development and prospective evaluations will be needed to substantiate the model’s performance and generalizability. We used a cluster of comorbidity codes to define GIB that did not differentiate between types and does not include all possible bleeding codes. Given visceral and non-visceral bleeds in the upper-, middle-, and lower-GI system can have differences in etiology, onset, treatment strategies, and outcomes,^31, 32, 53^ predictive models could be designed for specific GIB types. However, this would yield lower incidence rates and could hinder model performance. Our model can inherently account for collinearity and missingness, which is a strength. Nonetheless, the model determines associations and thereby predictors may not represent causal relationships and need to be interpreted within this context. There is always some give and take with regards to optimizing the sensitivity and specificity of any model, and our prediction model provides a more specific than sensitive prediction, which should also be considered in interpretation of results. A sufficient bleeding risk model may provide early detection of a HD patient’s ensuing GIB event in hopes that timely assessment and referral could limit avoidable hospitalization and the risk attributable to experiencing a major GIB.

Overall, we developed a machine learning model with suitable performance in classifying the 180-day risk of a GIB hospitalization for a HD patient. This model offers a potential method for early detection and prospective evaluations appear warranted. **Figure 3** shows a hypothetical workflow we propose for testing the model in clinical decision support. We suggest routine predictions of GIB risk on a quarterly basis with reporting after comprehensive labs that include total serum 25OH vitamin D. Reports could include binary classification (high vs low risk), or several risk classifications based on defined cutoff thresholds (e.g., >90%, ≤ 90% to >80%, ≤ 80% risk for GIB hospitalization). For each patient classified at a higher risk, we recommend the report should show at a minimum the top five predictors attributable to that patient’s prediction. Optimally, the reporting could be incorporated into everyday practice and considered at routine visits, just as lab values would be.

**Figure 3:**
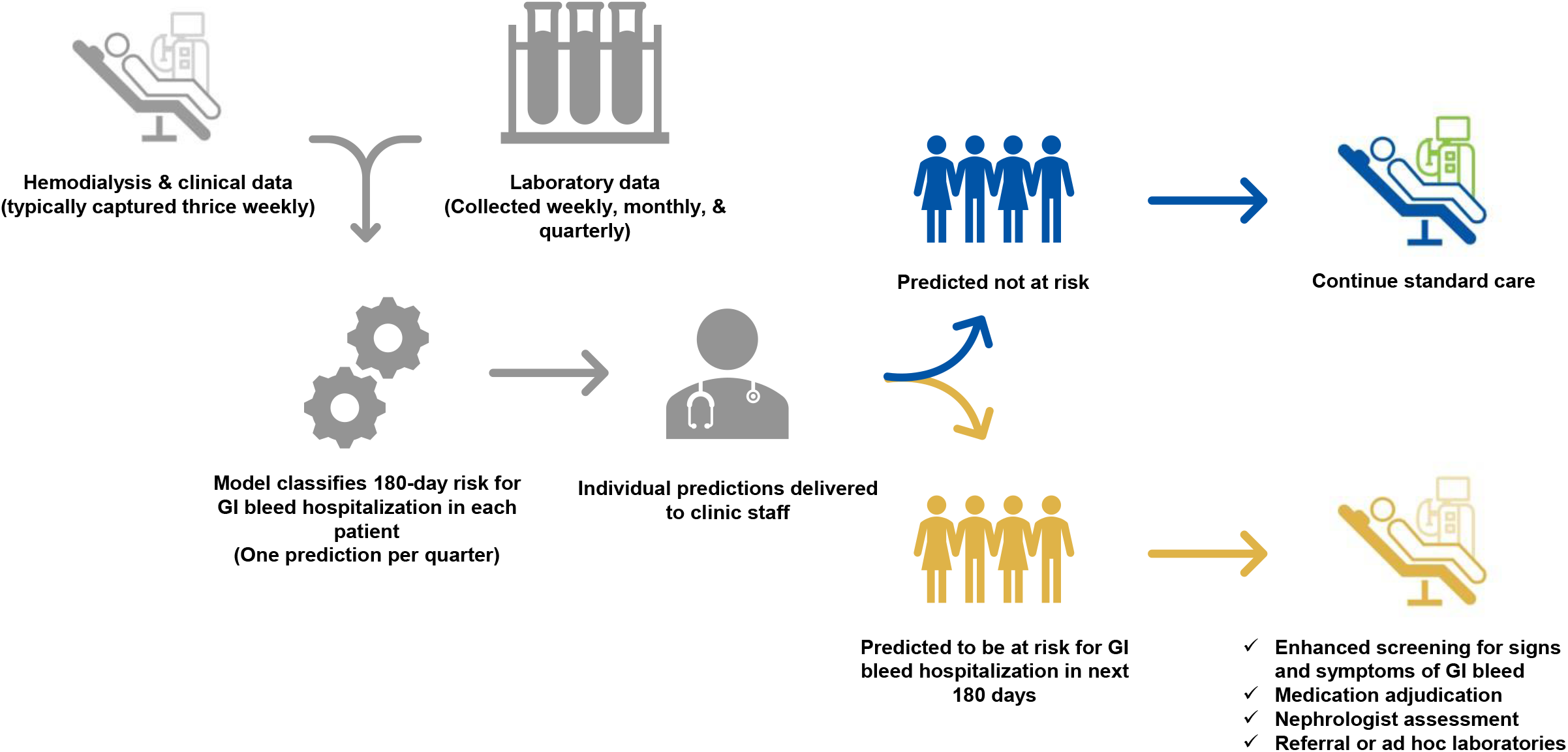
Hypothetical Workflow for Decision Support

## Data Availability

The dataset used in the presented analyses is not publicly available. The dataset was captured from a private electronic medical record system that is restricted to use by only authorized employees of Fresenius Medical Care. The dataset can be made available upon reasonable request to access the dataset, which would require an agreement to be established between Fresenius Medical Care and an external institution of any applicable requestor.

## Acknowledgements

We would like to acknowledge support provided by Diane M. Rondeau in scheduling project meetings and consultation provided from Caitlin Monaghan, PhD, Linda Ficociello, DSc, and Derek Blankenship, PhD, MBA in the design of the analysis, Katrin Köhler, MSc, MBA for administration of agreements in creation of the INSPIRE Core Group, and Carly Hahn Contino, MBA, PMP for being the initial INSPIRE project manager. Also, we express our sincere gratitude to the patient care teams at Fresenius Kidney Care in the United States who charted the real-world data used for this analysis during standard care, and most importantly directly provided kidney replacement therapy to improve the quality and quantity of life for the patients the overall nephrology community serves.

## References

1. Sood MM, Larkina M, Thumma JR, et al. Major bleeding events and risk stratification of antithrombotic agents in hemodialysis: results from the DOPPS. Kidney Int 2013; 84: 600–608.

2. Molnar AO, Bota SE, Garg AX, et al. The Risk of Major Hemorrhage with CKD. J Am Soc Nephrol 2016; 27: 2825–2832.

3. Lutz J, Menke J, Sollinger D, et al. Haemostasis in chronic kidney disease. Nephrol Dial Transplant 2014; 29: 29–40.

4. Garg R, Parikh MP, Chadalvada P, et al. Lower rates of endoscopy and higher mortality in end-stage renal disease patients with gastrointestinal bleeding: A propensity matched national study. J Gastroenterol Hepatol 2022; 37: 584–591.

5. Ishigami J, Grams ME, Naik RP, et al. Chronic Kidney Disease and Risk for Gastrointestinal Bleeding in the Community: The Atherosclerosis Risk in Communities (ARIC) Study. Clin J Am Soc Nephrol 2016; 11: 1735–1743.

6. Yang JY, Lee TC, Montez-Rath ME, et al. Trends in acute nonvariceal upper gastrointestinal bleeding in dialysis patients. J Am Soc Nephrol 2012; 23: 495–506.

7. Holden RM, Harman GJ, Wang M, et al. Major bleeding in hemodialysis patients. Clin J Am Soc Nephrol 2008; 3: 105–110.

8. Nopp S, Spielvogel C, Schmaldienst S, et al. Bleeding risk assessment in end-stage kidney disease: validation of existing risk scores and evaluation of a machine learning-based approach. Thromb Haemost 2022.

9. Niikura R, Aoki T, Kojima T, et al. Natural history of upper and lower gastrointestinal bleeding in hemodialysis patients: A dual-center long-term cohort study. J Gastroenterol Hepatol 2021; 36: 112–117.

10. van Eck van der Sluijs A, Abrahams AC, Rookmaaker MB, et al. Bleeding risk of haemodialysis and peritoneal dialysis patients. Nephrol Dial Transplant 2021; 36: 170–175.

11. Trivedi H, Yang J, Szabo A. Gastrointestinal bleeding in patients on long-term dialysis. J Nephrol 2015; 28: 235–243.

12. Blatchford O, Murray WR, Blatchford M. A risk score to predict need for treatment for upper-gastrointestinal haemorrhage. Lancet 2000; 356: 1318–1321.

13. Pisters R, Lane DA, Nieuwlaat R, et al. A novel user-friendly score (HAS-BLED) to assess 1-year risk of major bleeding in patients with atrial fibrillation: the Euro Heart Survey. Chest 2010; 138: 1093–1100.

14. Singer DE, Chang Y, Borowsky LH, et al. A new risk scheme to predict ischemic stroke and other thromboembolism in atrial fibrillation: the ATRIA study stroke risk score. J Am Heart Assoc 2013; 2: e000250.

15. Gage BF, Yan Y, Milligan PE, et al. Clinical classification schemes for predicting hemorrhage: results from the National Registry of Atrial Fibrillation (NRAF). Am Heart J 2006; 151: 713–719.

16. O’Brien EC, Simon DN, Thomas LE, et al. The ORBIT bleeding score: a simple bedside score to assess bleeding risk in atrial fibrillation. Eur Heart J 2015; 36: 3258–3264.

17. Ocak G, Ramspek C, Rookmaaker MB, et al. Performance of bleeding risk scores in dialysis patients. Nephrol Dial Transplant 2019; 34: 1223–1231.

18. Satilmis D, Yavuz BG, Guven O, et al. The effectiveness of Glasgow-Blatchford Score in early risk assessment of hemodialysis patients. Intern Emerg Med 2021.

19. Chaudhuri S, Long A, Zhang H, et al. Artificial intelligence enabled applications in kidney disease. Semin Dial 2021; 34: 5–16.

20. Stennett A, Mysayphonh C, Kovacevic T, Larkin JW, Guedes M, Moraes TP, Maddux FW, Chatoth D, Pecoits-Filho R, Hymes J. Provider evaluates methoxy polyethylene glycol-epoetin beta in peritoneal dialysis. Nephrol News Issues 2020; 34: https://www.healio.com/news/nephrology/20200909/provider-evaluates-methoxy-polyethylene-glycolepoetin-beta-in-peritoneal-dialysis.

21. Fadem SZ, Al-Saghir F, Zollner G, et al. Converting hemodialysis patients from intravenous paricalcitol to intravenous doxercalciferol - a dose equivalency and titration study. Clin Nephrol 2008; 70: 319–324.

22. National Kidney F. K/DOQI clinical practice guidelines for bone metabolism and disease in chronic kidney disease. Am J Kidney Dis 2003; 42: S1–201.

23. National Drug Code Directory. United States Food and Drug Administration: https://www.fda.gov/drugs/drug-approvals-and-databases/national-drug-code-directory.

24. Chen T, Guestrin C: XGBoost: A Scalable Tree Boosting System. In Proceedings of the 22nd ACM SIGKDD International Conference on Knowledge Discovery and Data Mining, San Francisco, California, USA, Association for Computing Machinery, 2016, pp 785–794

25. Shapley LS. “A Value for n-Person Games,” In: H. W. Kuhn and A. W. Tucker, Eds., Contributions to the Theory of Games II. Annals of Mathematics Studies, Princeton University Press, Princeton 1953; 28: 307–317.

26. Štrumbelj E, Kononenko I. Explaining prediction models and individual predictions with feature contributions. J Knowledge and Information Systems 2013; 41: 647–665.

27. Lundberg S, Lee SI. “A Unified Approach to Interpreting Model Predictions.” In: I. Guyon, U. V. Luxburg, S. Bengio, H. Wallach, R. Fergus, S. Vishwanathan and R. Garnett, Eds., Advances in Neural Information Processing Systems 30. Curran Associates, Inc 2017: 4765–4774.

28. Lundberg SM, Erion G, Chen H, et al. From local explanations to global understanding with explainable AI for trees. Nature Machine Intelligence 2020; 2: 56–67.

29. Wasse H, Gillen DL, Ball AM, et al. Risk factors for upper gastrointestinal bleeding among end-stage renal disease patients. Kidney Int 2003; 64: 1455–1461.

30. Tokar JL, Higa JT. Acute Gastrointestinal Bleeding. Ann Intern Med 2022; 175: ITC17–ITC32.

31. Laine L, Barkun AN, Saltzman JR, et al. ACG Clinical Guideline: Upper Gastrointestinal and Ulcer Bleeding. Am J Gastroenterol 2021; 116: 899–917.

32. Strate LL, Gralnek IM. ACG Clinical Guideline: Management of Patients With Acute Lower Gastrointestinal Bleeding. Am J Gastroenterol 2016; 111: 755.

33. Cooper GS, Kou TD, Wong RC. Outpatient management of nonvariceal upper gastrointestinal hemorrhage: unexpected mortality in Medicare beneficiaries. Gastroenterology 2009; 136: 108–114.

34. Laeeq SM, Tasneem AA, Hanif FM, et al. Upper Gastrointestinal Bleeding in Patients with End Stage Renal Disease: Causes, Characteristics and Factors Associated with Need for Endoscopic Therapeutic Intervention. J Transl Int Med 2017; 5: 106–111.

35. Sotoudehmanesh R, Ali Asgari A, Ansari R, et al. Endoscopic findings in end-stage renal disease. Endoscopy 2003; 35: 502–505.

36. Deshmukh F, Merchant SS. Explainable Machine Learning Model for Predicting GI Bleed Mortality in the Intensive Care Unit. Am J Gastroenterol 2020; 115: 1657–1668.

37. Aoki T, Yamada A, Nagata N, et al. External validation of the NOBLADS score, a risk scoring system for severe acute lower gastrointestinal bleeding. PLoS One 2018; 13: e0196514.

38. Oakland K, Jairath V, Uberoi R, et al. Derivation and validation of a novel risk score for safe discharge after acute lower gastrointestinal bleeding: a modelling study. Lancet Gastroenterol Hepatol 2017; 2: 635–643.

39. Tapaskar N, Jones B, Mei S, et al. Comparison of clinical prediction tools and identification of risk factors for adverse outcomes in acute lower GI bleeding. Gastrointest Endosc 2019; 89: 1005–1013 e1002.

40. Benedeto-Stojanov D, Bjelakovic M, Stojanov D, et al. Prediction of in-hospital mortality after acute upper gastrointestinal bleeding: cross-validation of several risk scoring systems. J Int Med Res 2022; 50: 3000605221086442.

41. Keskin U, Basat S. The effect of vitamin D levels on gastrointestinal bleeding in patients with warfarin therapy. Blood Coagul Fibrinolysis 2019; 30: 331–336.

42. Vitamin D: Fact Sheet for Health Professionals. National Institutes of Health: https://ods.od.nih.gov/factsheets/VitaminD-HealthProfessional/#en1.

43. Kidney Disease: Improving Global Outcomes CKDMBDUWG. KDIGO 2017 Clinical Practice Guideline Update for the Diagnosis, Evaluation, Prevention, and Treatment of Chronic Kidney Disease-Mineral and Bone Disorder (CKD-MBD). Kidney Int Suppl (2011) 2017; 7: 1–59.

44. Larkin JW, Jiao Y, Lama SK, Chaudhuri S, Willetts J, Winter A, Stauss-Grabo M, Usvyat LA, Hymes JL, Maddux FW, Stenvinkel P, Floege J. Higher 25-Hydroxyvitamin D Associates With Gastrointestinal Bleeding Events (Abstract TH-PO837). J Am Soc Nephrol 2022; 33: 287.

45. Mohammad S, Mishra A, Ashraf MZ. Emerging Role of Vitamin D and its Associated Molecules in Pathways Related to Pathogenesis of Thrombosis. Biomolecules 2019; 9.

46. Puri S, Bansal DD, Uskokovic MR, et al. Induction of tissue plasminogen activator secretion from rat heart microvascular cells by fM 1,25(OH)(2)D(3). Am J Physiol Endocrinol Metab 2000; 278: E293–301.

47. Fukumoto S, Allan EH, Martin TJ. Regulation of plasminogen activator inhibitor-1 (PAI-1) expression by 1,25-dihydroxyvitamin D-3 in normal and malignant rat osteoblasts. Biochim Biophys Acta 1994; 1201: 223–228.

48. Barbosa EM, Nonogaki S, Katayama ML, et al. Vitamin D3 modulation of plasminogen activator inhibitor type-1 in human breast carcinomas under organ culture. Virchows Arch 2004; 444: 175–182.

49. Koyama T, Shibakura M, Ohsawa M, et al. Anticoagulant effects of 1alpha,25-dihydroxyvitamin D3 on human myelogenous leukemia cells and monocytes. Blood 1998; 92: 160–167.

50. Aihara K, Azuma H, Akaike M, et al. Disruption of nuclear vitamin D receptor gene causes enhanced thrombogenicity in mice. J Biol Chem 2004; 279: 35798–35802.

51. Lindqvist PG, Epstein E, Olsson H. Does an active sun exposure habit lower the risk of venous thrombotic events? A D-lightful hypothesis. J Thromb Haemost 2009; 7: 605–610.

52. Beer TM, Venner PM, Ryan CW, et al. High dose calcitriol may reduce thrombosis in cancer patients. Br J Haematol 2006; 135: 392–394.

53. Muftah M, Mulki R, Dhere T, et al. Diagnostic and therapeutic considerations for obscure gastrointestinal bleeding in patients with chronic kidney disease. Ann Gastroenterol 2019; 32: 113–123.

