## supplementary for "Prediction of Gastrointestinal Bleeding Hospitalization in Hemodialysis"

### Supplementary Materials:

**SUPPLEMENTARY TABLE 1: EXPOSURE VARIABLE DESCRIPTIONS CONSIDERED IN INITIAL MODEL DEVELOPMENT**

| Parameters | Distinct Variables | All Input Variables | Description |
|---|---|---|---|
| **Demographics** | | | |
| Age | 1 | 1 | Age (years) |
| Sex | 1 | 1 | Male (versus Female) |
| Race | 5 | 5 | Asian, Black, White, other, unknown |
| Ethnicity | 3 | 3 | Hispanic, Not Hispanic, unknown |
| Height | 1 | 1 | Centimeters tall |
| Dialysis vintage | 1 | 1 | Years on chronic dialysis |
| Marital status | 4 | 4 | Single, Married/partner/union, Divorced/separated/widowed, unknown |
| Dialysis access | 2 | 2 | Catheter, Fistula/Graft |
| **Comorbidities** | | | |
| Self-reported energy | 1 | 1 | KDQOL-36 score for question 10: “Did you have a lot of energy?” |
| Total chronic comorbidity duration | 10 | 10 | Anemias, Cancer other than skin, Cerebrovascular disease, Chronic obstructive pulmonary disease, Congestive heart failure, Diabetes, Hepatitis, Hyperparathyroidism, Ischemic heart disease, Peripheral vascular/arterial disease (years since start) ◊ |
| Time since acute morbidity | 5 | 5 | Cardiac arrest, Cardiac dysrhythmias, Digestive disorders other than GI bleed, GI bleed, Infection (time since most recent event/active status) ◊ |
| **Environmental** | | | |
| Season | 4 | 4 | Winter (Dec-Feb), spring (Mar-May), summer (Jun-Aug), autumn (Sep-Nov) |
| **Laboratories** | | | |
| Cell blood counts | 8 | 35 | Hemoglobin (weekly), white blood cells, neutrophils, lymphocytes, platelets, monocytes, eosinophils, basophils (monthly), hemoglobin A1C (if has diabetes) (quarterly) |
| Chemistry | 12 | 48 | Albumin, calcium, corrected calcium, chloride, creatinine, bicarbonate, phosphate, potassium, sodium, blood urea nitrogen, blood urea nitrogen to creatinine ratio, urea reduction ratio (monthly) |
| Bone factors | 2 | 6 | Intact parathyroid hormone, total 25OH vitamin D (quarterly) |
| Iron indices | 2 | 6 | Transferrin saturation, ferritin (quarterly) |
| Liver function tests | 2 | 6 | Alkaline phosphatase, alanine transaminase (quarterly) |
| Inflammatory markers | 2 | 6 | Neutrophil to lymphocyte ratio (monthly), C-reactive protein (*ad hoc*) |
| Clot factors | 2 | 4 | Prothrombin time, international normalized ratio (if on warfarin) (*ad hoc*) |
| B vitamins | 2 | 4 | Folic acid (annual), vitamin B12 (on admission to outpatient HD and *ad hoc*) |
| **HD Treatment Data** | | | |
| Vital signs | 12 | 48 | Standing and sitting systolic and diastolic blood pressure, sitting heart rate, temperature (pre-HD & post-HD) |
| Weights | 5 | 20 | Weights last HD (pre-HD & post-HD), estimated dry weight (EDW), removed weight as percent of EDW, removed weight as percent of target to remove |
| Dialysis delivery | 6 | 24 | Treatment time, KECN (effective conductivity clearance of sodium), online clearance Kt/V, Qb, Qd, saline administration |

| | | | |
|---|---|---|---|
| Shortened HD session | 14 | 56 | Ended treatment early: against medical advice, patient request, physician request, patient late, complication (clotted access, poor flows, hypotension, technical difficulty, system problem), emergency, hospitalization, unexpected, other, unknown |
| Rescheduled HD session | 5 | 7 | Days since start/end last rescheduled HD, days between rescheduled HD to next session, ≥1 rescheduled HD in last 180 days, number of rescheduled HD (last 30, 90, 180 days) |
| Missed HD treatments | 5 | 7 | Days since start/end last missed HD, days between missed HD to next session, ≥1 missed HD in last 180 days, number of missed HD (last 30, 90, 180 days) |
| **Medications** | | | |
| In-center medications | 13 | 52 | Systemic heparin, heparin catheter lock IV vitamin D, oral vitamin D (calcitriol, paricalcitol, or ergocalciferol), calcimimetic (etelcalcetide, cinacalcet), erythropoietin stimulating agents, IV iron (iron sucrose, sodium ferric gluconate, or ferumoxytol), 4% sodium citrate (dose of medication) |
| Home medications | 5 | 5 | Proton pump inhibitors, warfarin, apixaban, fondaparinux, antiplatelets (e.g. platelet aggregation inhibitors) (use of medication) ◊◊ |
| **Events** | | | |
| All cause hospitalizations/events | 6 | 8 | Days since start last hospitalization, ≥1 hospitalization in last 180 days, length of stay (days) for last hospitalization, number of hospitalizations (last 30, 90, 180 days), emergency room visit in last 180 days, temporary transfer outside provider in last 180 days |
| GI bleed hospitalizations | 4 | 6 | Days since start last GI bleed hospitalization, ≥1 GI bleed hospitalization in last 180 days, length of stay (days) for last GI bleed hospitalization, number of GI bleed hospitalizations (last 30, 90, 180 days) |

Comorbidities: ◊ ICD10 groupings for all comorbidities available upon request

Laboratories, HD treatment data, in-center medications: Model will consider most recent value for each distinct variable, as well as the mean values in the prior 7, 30, 90, and/or 180 days for each distinct variable as deemed appropriate considering data frequency/availability (represented in all variables column of table).

Medications: ◊◊ Pharmacologic classes defined by the United States Food and Drug Administration's National Drug Code Directory; no patients were administered edoxaban or rivaroxaban and these medications were omitted from the model.

Events: Model will consider most recent value for each distinct variable.

**SUPPLEMENTARY TABLE 2: DEFINITION OF TRUE AND FALSE POSITIVE AND NEGATIVE PREDICTIONS CLASSIFIED BY THE MODEL**

| | |
|---|---|
| **True positives** | Patients classified as having a GI bleed event by the model who are in the group with a GI bleed event |
| **False positives** | Patients classified as having a GI bleed event by the model who are in the group without a GI bleed |
| **True negatives** | Patients classified as not having a GI bleed event by the model who are in the group without a GI bleed |
| **False negatives** | Patients classified as not having a GI bleed event by the model who are in the group with a GI bleed event |

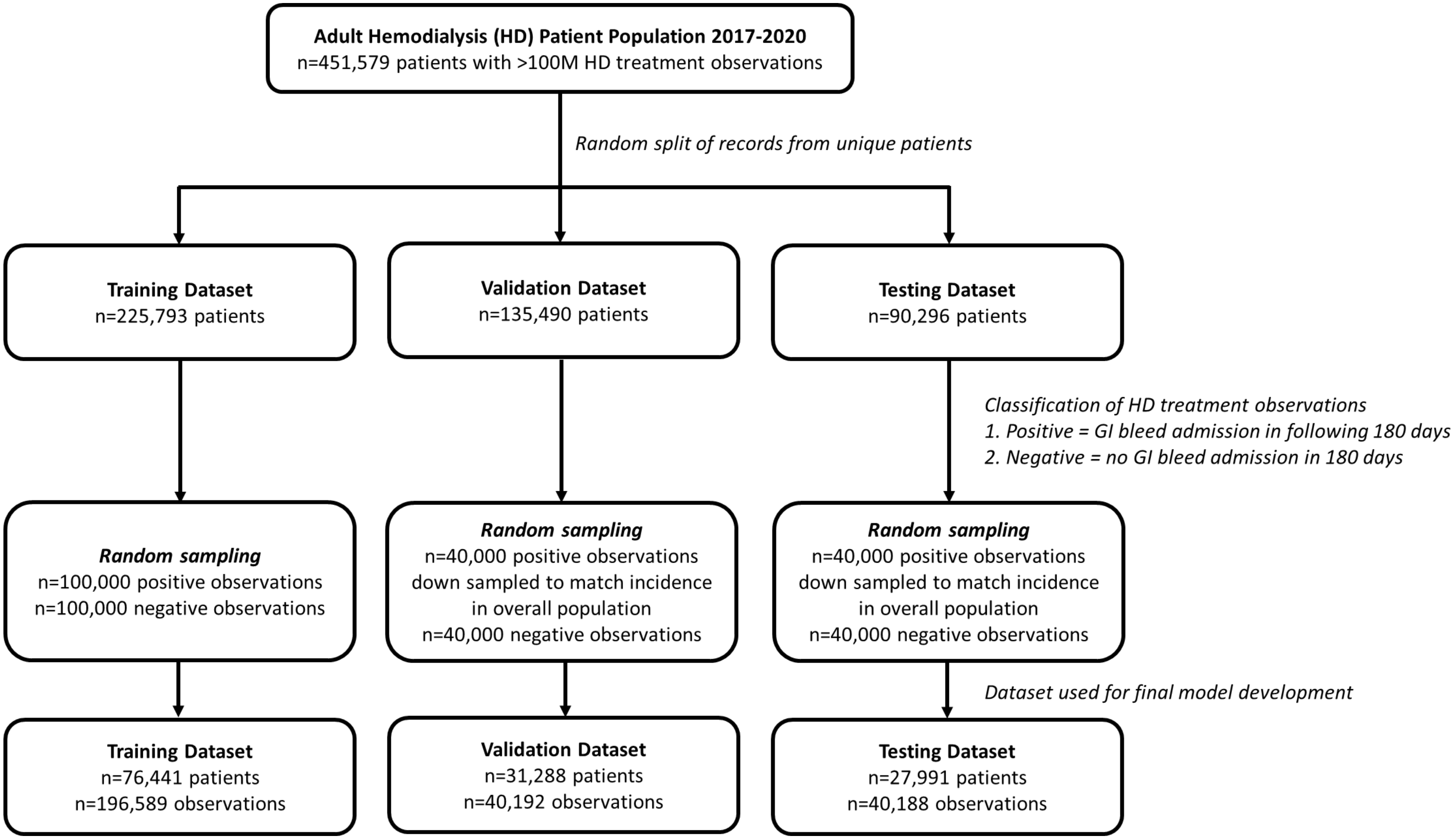


**Supplementary Figure 1:** Selection of data for model development

**Appendices:**

**Appendix A:**

**INSPIRE CORE GROUP MEMBERS, AFFILIATIONS, AND ROLES**

| Member | Institution | Role |
|---|---|---|
| Jürgen Floege, MD | University Hospital Aachen | Medical subject matter expert |
| Peter Stenvinkel, MD, PhD, FASN | Karolinska Institutet | Medical subject matter expert |
| David C. Wheeler, MB, CHB, MD, FRCP | University College London | Medical subject matter expert |
| Franklin W. Maddux, MD, FACP | FME, GMO, Europe & North America | Medical subject matter expert |
| Jeffrey L. Hymes, MD | FME, GMO, North America | Medical subject matter expert |
| Nwamaka Eneanya, MD, MPH, FASN | FME, GMO, North America | Medical subject matter expert |
| Len A. Usvyat, PhD | FME, GMO, North America | Global data management and analytics |
| Manuela Stauss-Grabo, PhD | FME, GMO, Europe | Global biomedical evidence generation |
| Anke Winter, MD, MSc | FME, GMO, Europe | Data management and analytics (International outside United States) |
| Sheetal Chaudhuri, MS | FME, GMO, North America | Data management and analytics (United States) |
| John W. Larkin, PhD, MSc, CCRC | FME, GMO, North America | Collaborations, investigations, and publications |
| Justin Zimbelman, MBA | FME, North America | Project manager |
